# Health-Related Quality of Life Among Critical Ill Patients After Move Out of ICU - A Systematic Review Protocol

**DOI:** 10.1101/2022.06.23.22276815

**Authors:** Yao Li, Dan Fang, Qiao Wu

## Abstract

**Introduction:** the impact of critical illness is profound on patients resulting in physical, mental and social consequences and a poor health-related quality of life (HRQOL). Several studies investigated HRQOL among patients after moving out of the Intensive care unit (ICU). We will propose a systematic review of these studies.

**Method:** We will search PubMed, Web of Science, EMBASE, CINAHL (Cumulative Index to Nursing and Allied Health Literature), Cochrane Library and open grey paper that reported HRQOL of ICU survivors from January 2012 to January 2022 in the English language. We will exact HRQOL data. Risk of bias will use the QUADAS-2 tool. The strength of results depends on the quality of studies.

**Strengths and limitations of this study:** This study will focus on general ICU survivors and make sure our results are generalizable

The information about HRQOL is updated, and the follow-up period was extended. We will focus on recently ten-year studies. We will be glad to see whether the HRQOL improved.

This study will analyze the factors associated with HRQOL after patients moved out of ICU.

This study will not perform a meta-analysis due to the heterogeneous population.

**Ethics and dissemination:** We do not require ethical approval because our review will be based on published material.

**Trial registration number:** PROSPERO CRD 42022304279

## INTRODUCTION

### Rationale

With improved intensive care medicine, most critically ill patients can survive critical conditions and move out of the Intensive care unit (ICU). According to Society of vital care medicine statistics, over five million patients are admitted to ICU annually in the USA, and over 80% of patients could be discharged from the ICU (SCCM, 2020). Meanwhile, these patients were called ICU survivors. They benefited from advanced intensive care while suffering the sequela of critical illness. Over half of ICU survivors suffered different degrees of physical, mental, and cognitive impairments called post-ICU syndrome (PICS) and poor HRQOL after discharge from ICU (2,3). HRQOL was a perception of the life of patients affected by critical illness and ICU experience (4). ICU survivors needed to reflect on functioning outcomes from patients’ perspectives. Poor HRQOL among ICU survivors becomes a significant concern linked with higher mortality, financial burdens, and family caregivers’ burdens (5–7). In the past tens of years, poor HRQOL among ICU survivors addressed attention from researchers, which led to an increasing number of HRQOL research. Thus, we will conduct a systematic review to analyze and summarise the results.

We will use the valid instrument SF 36 to measure HRQOL. Due to the heterogeneity of ICU survivors, generic instruments such as SF family tools include SF36, SF 12, SF 20 and EQ-5D. However, only SF 36 had dimensions scores that reflect HRQOL comprehensively. Therefore, this study analyses the results that used valid interments of SF 36 (version 1 and version 2). This review will review the existing literature on HRQOL in SF 36 instrument after patients moved out of ICU and the measurement time. We also measured HRQOL compared to the general population, which explains HRQOL changes with time following ICU discharge. We will identify factors associated with HRQOL. We are interested in determining the length of follow-up and how many patients are lost to follow-up. We will provide comprehensive and detailed descriptive information for future reference in long-term outcome research for general ICU survivors.

### Objective

In this study, our primary objective is to explore HRQOL among patients discharged from ICU compared with the general population. Our second outcome is to examine the change of HRQOL with time and examine factors that affected HRQOL in studies and follow-up duration.

## Methods

This protocol followed (PRISMA-P) guidelines. We will conduct a systematic review of literature on HRQOL among patients discharged from ICU. The language is English.

### Information sources

we searched PubMed, Web of Science, EMBASE, Cochrane Library, CINAHL, and open grey paper that reported HRQOL of ICU survivors from January 2012 to January 2022 in English.

### Search strategy

Our search strategy is (“critically ill patients” OR “critical ill survivor” OR “ICU patients” OR “ICU survivor”) AND (“Health-related quality of life” OR “quality of life” OR “HRQOL” OR “QQL” OR “long-term outcome”). We reviewed the reference list of primary studies included in the review and the reference lists of relevant, previously published reviews. We also investigated why the initial search strategy had not been discovered. A new ‘key phrase’ will be used if it is necessary.

### Eligible criteria

From 2011 to 2021, we will look at prospective cohort studies, retrospective cohort studies, and cross-sectional studies published in English.

In a diagram adapted from the PRISMA guidelines, the inclusion of studies in each stage of the systematic review will be shown (8)

### Report inclusion criteria

1. Adult ICU survivors: age over 18
2. Original research
3. Quality of life assessed at least discharge for ICU three months
4. Measurement and reporting of specific QOL domains at baseline and follow-up.
5. Compared with the age-adjusted population
6. General ICU settings (medical/surgical/mixed)
7. Valid measurement SF 36 (Version one / version two)
8. English language

### Exclude criteria

1. Only investigated precisely procedure or event, for example, post-cardiac arrest, COVID-19, ECOM patients
2. Specific age population, For instance, age over 65
3. Only in particular diseases or one disease, For example, trauma.
4. Experimental studies without a control group.
5. We would choose a study with the longest follow-up time if the studies reported the same patients.

### Data management

we will store identified citations and electronic text to store a reference manager programme.

### Selection process

After the search, two reviewers will independently screen the titles and abstracts that identify potential studies. The process will follow Prisma-P guideline 2020 (figure 1). Two researchers will remove abstracts that do not match the eligibility criteria. If there is no consensus, we will have a discussion. If necessary, the third researcher was consulted to make the final decision.

**Figure one:**
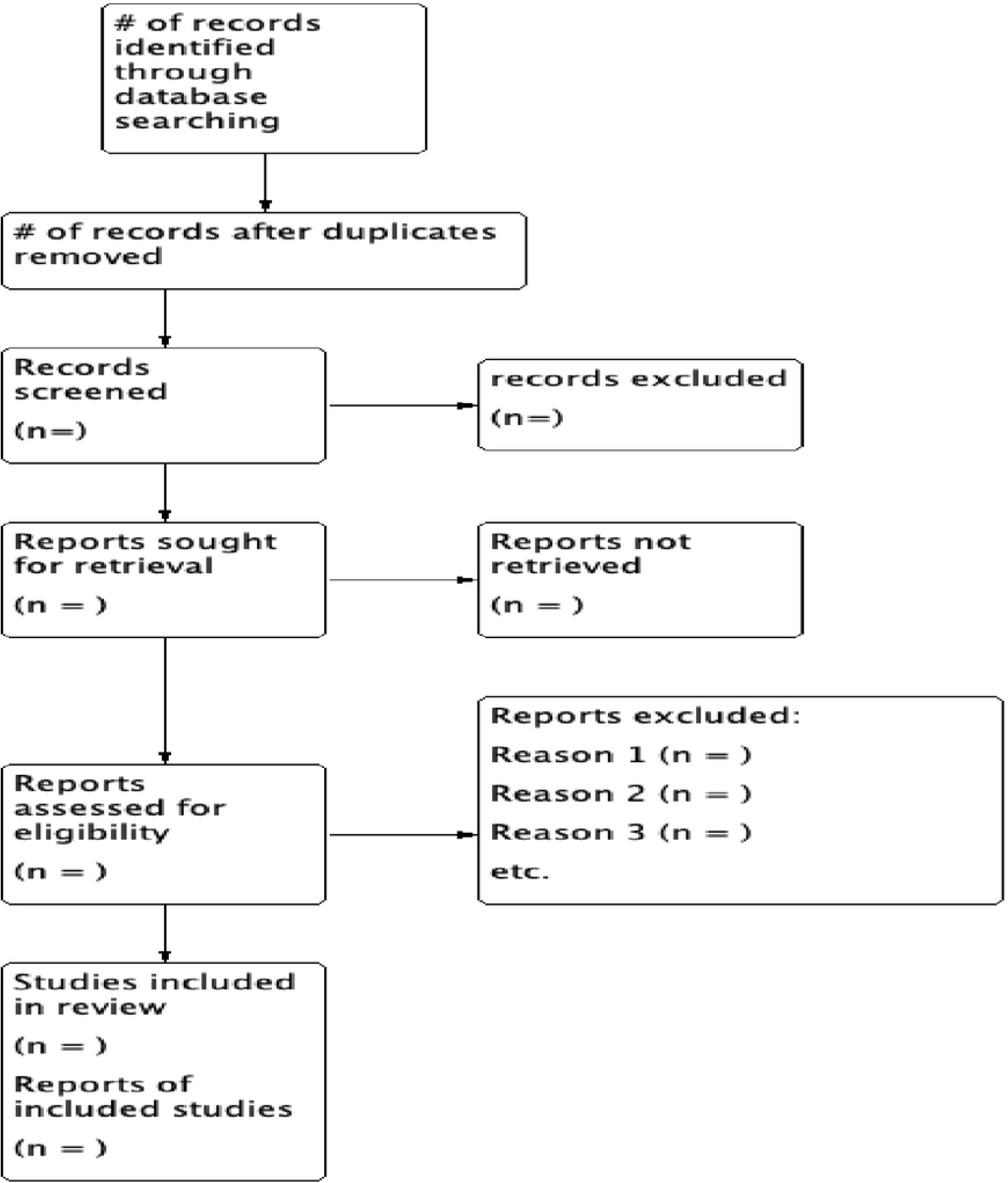
The selection flow diagram, adapted from PRISMA statement 2020 (8).

Two researchers will independently carry out a final screening process of the full-text articles to select articles that reach our eligibility criteria. Again, we will record the reason for the exclusion. The inclusion of papers in each stage of the systematic review will be shown in a diagram adapted from the PRISMA statement(8)

## Data extraction

Two review authors will extract data from the full-text article and online supplementary data and will be performed as a data extraction table. It will not be included if a consensus of studies cannot be reached. In case of unclear information, we will contact the authors and exclude ambiguous data if this is unsuccessful in reaching the author.

We will extract the following information:

1. Citation ’2 Study country of origin
2. Study design
3. Year of publication
4. Type of ICU
5. Patient type: general/sepsis/respiratory.
6. The number of patients was screened.
7. The total number of patients who were alive at the start of the study.
8. Successful Follow-up rate
9. Follow-up time points
10. Proxy ICU HRQOL assess
11. Data on HRQOL gleaned from studies and online resources
12. HRQOL relative to the age-adjusted average population
13. Previous health/pre-existing disease
14. The trend in HRQOL post-discharge.
15. Factors affecting HRQOL in post-discharge

### Synthesis of data and data analysis

We will describe ICU survivors’ health-related quality of life and subgroups (sepsis, pulmonary, general) (tabular). We will describe secondary outcomes (factors that affect HRQOL, length follow-up in tabular or graphical form.

### Bias assessment

We will set the papers for applicability and bias using the QUADAS-2 tool (9). Due to the heterogeneity of study designs, we have no plans to conduct a meta-analysis of the study findings or assess meta-biases statistically.

### Assessment of the strength of the conclusion

The quality of studies determines the strength of the conclusion.

Indeed, previous four studies had conducted HRQOL research. However, most studies retained the studies for 20 years that cannot accurately reflect HRQOL change among ICU survivors (Dowdy et al., 2005; Elliott, 1999; Gerth et al., 2019; Oeyen et al., 2010). Meanwhile, previous system reviews mostly retained studies in developed countries (10,13,14). In recent years, poor HRQOL among ICUs addressed much more attention in developed countries (15,16). Thus, previous studies could not reflect HRQOL among ICU survivors comprehensively. Elliott and colleagues 1999 reviewed 31 studies, but fewer studies used valid instruments (11). Dowdy et al. (2005) rexamined21 studies that included 7320 patients and found ICU survivors suffered poor HRQOL; meanwhile, age and severity of illness were the associated factors. Oeyen et al. (2010) retained 53 studies that found that HRQOL was not ideal compared with the norm population. The impact factors of HRQOL were severity of illness, comorbidity, age, and gender. However, they retained studies on specific diseases or events such as cardiac arrest and acute pancreatitis. A newly published systematic review from Gerth et al. (2019) included 48 studies from 2000 to 2015. They found that ICU survivors had poor HRQOL compared to the sex-matched population and improved with time. The most extended follow-up duration was 60 months. However, they did not analyze what factors affect HRQOL (Gerth et al., 2019). With increased awareness of the long-term outcome of ICU, Haug and colleagues conducted a systematic review in a subgroup of Sepsis, major trauma, and severe burn injury. They found that HRQOL among these groups showed similarities to HRQOL.

Nevertheless, it may be less generable to represent general ICU survivors’ health status (17). This research will concentrate on the foundations of previous research. We looked at studies from the last ten years. Rather than focusing on specific events, we aimed for general ICU admission. Studies that used reliable instruments will be kept. We will look at the factors that influence HRQOL. To date, the follow-up duration has been extended. We would like to see whether the HRQOL would improve with time.

## Data Availability

No datasets were generated or analysed during the current study. All relevant data from this study will be made available upon study completion.

## Author Contributions

Yao Li designed a systematic review and wrote this protocol. Dan Fang and Qiao Wu screened articles, extracted data, assessed bias and processed data independently. During the study, any disagreement will be arbitrated by Yao Li. All authors agree with the publication of this protocol.

The authors have no conflict of interest.

## Funding

no funding

## Acknowledgements

None

## Conflict interests

None declared.

## Notes

### Competing Interest Statement

The authors have declared no competing interest.

### Clinical Protocols

N?A

### Funding Statement

The author(s) received no specific funding for this work.

### Author Declarations

We do not require ethical approval because our review will be based on published material.

